# Closed Doors: Predictors of Stress, Anxiety, Depression, and PTSD During the Onset of COVID-19 Pandemic in Brazil

**DOI:** 10.1101/2021.08.18.21262061

**Authors:** Vitor Crestani Calegaro, Luis Francisco Ramos-Lima, Mauricio Scopel Hoffmann, Gustavo Zoratto, Natália Kerber, Fernanda Coloniese Dala Costa, Vitor Daniel Picinin, Julia Köchler, Leonardo Rodrigues, Luisa Maciel, Luiza Elizabete Braun, Fernando Leite Girardi, Gabriel Olerich Cecatto, Leopoldo Pompeo Weber, Bruna Fragoso Rodrigues, Alessandra Naimaier Bertolazi, Juliana Motta de Oliveira, Bianca Lorenzi Negretto, Andrea Feijó de Mello

## Abstract

**Background:** The rise of mental health problems in the population directly or indirectly by the COVID-19 pandemic is a major concern. The aim of this study was to investigate and compare independent predictors of symptoms of stress, anxiety, depression, and post-traumatic stress disorder (PTSD) in Brazilians, one month after the implementation of measures of social distancing.

**Methods:** It was a cross-sectional study, performed through a web-based survey. Depression, Anxiety, and Stress Scale (DASS-21) and PTSD Checklist for DSM-5 (PCL-5) were the outcomes. Data were gathered regarding demographics, social distancing, economic problems, exposure to the news of the pandemic, psychiatric history, sleep disturbances, traumatic situations, and substance use. The Alcohol Use Disorders Identification Test - Consumption (AUDIT-C) was also included. Predictors of symptoms were investigated through hierarchical multiple linear regression.

**Result:** Of a sample of 3,587 participants, approximately two-thirds considered that their mental health worsened after the beginning of the social restriction measures. The most important predictors of the symptoms investigated were the intensity of the distress related to pandemic news, younger age, current psychiatric diagnosis, trouble sleeping, emotional abuse or violence, and economic problems.

**Limitations:** The convenience sample assessed online may have limited external validity. It does not represent the northern regions of the country and most participants was white wealthier females.

**Conclusions:** These results confirm the hypothesis that a pandemic would have important impacts on the mental health of the population and indicate the level of distress related to the media as an important predictor of psychological suffering.

**Highlights:**

- Distress triggered by news was the main predictor of psychological symptoms
- Sleeping problems were strong indicators of mental health problems
- People with ongoing psychiatric disorders are especially vulnerable
- Measures to prevent interpersonal trauma and financial loss are crucial
- Young people may experience great suffering at the onset of the pandemic

## 1. Introduction

The emergence and rapid spread of COVID-19 with the potential to cause death in elder, adults, and even children, and substantial socioeconomic disruption, prompted health authorities to call for rapid measures (Holmes et al., 2020)(Kadir, 2020). Highly contagious infectious diseases, such as coronavirus disease 2019 (COVID-19), result in psychological distress to the population directly or indirectly affected. Stress may raise concomitantly with the expansion of the outbreak, which can trigger anxiety, depressive, and post-traumatic stress symptoms, especially when individuals experience the possibility of death - either in themselves or in loved ones (Sun et al., 2020). In addition, social restriction measures imposed during pandemic may result in several negative consequences to the mental health, producing stress overload. High rates of symptoms of stress, anxiety, depression, and post-traumatic stress disorder have been reported in the general population of low, middle, and high-income countries during pandemic (Xiong et al., 2020). All are affected, in a small or large extent, in a way to a possible second pandemic: of mental disorders (Choi et al., 2020).

The world population is facing new cultural and social rules, such as the regular use of facemasks and physical distancing. Mass quarantine was adopted worldwide in several contexts. Quarantine is conceptualized as the separation and restriction of movement in people who were exposed to a contagious disease to see if they become sick, aiming to reduce the risk of infecting others (CDC, 2019). Duration of quarantine, fear of infection, frustration, boredom, inadequate supplies and information along with feelings of helplessness and the loss of a sense of safety, security and financial stability evokes an increasingly familiar shudder of mistrust of others, avoidance, and withdrawal from everyday activities (Brooks et al., 2020). Social distancing associated with quarantine can be the catalyst for many mental health issues, even in previously healthy people (Usher et al., 2020).

Despite of the necessary measures to contain the rapid spread of the contagion, governmental focus has been fighting against infection, but the raising of mental disorders are often neglected (Ornell et al., 2020). In Brazil, the pre-pandemic prevalence of mental disorders was 50% higher than the global prevalence and, specifically for substance use and anxiety disorders, the prevalence was twice as high (Global Burden of Disease Collaborative Network, 2019). This continental country has the sixth higher population of the world, characterized by cultural heterogeneity and socioeconomical inequity. Like other low and middle-income countries, with low coverage of mental health care, the raise of mental disorders and lack of treatment may achieve epidemic proportions. Since posttraumatic stress disorder (PTSD), major depression and anxiety disorders have the potential to grow during pandemic, monitoring the mental health in the population is imperative to plan actions on prevention, health promotion, and treatment.

In this context, we developed the COVIDPsiq study (www.covidpsiq.org) to monitor the evolution of stress, depression, anxiety, and PTSD symptoms in Brazilians during the pandemic (www.covidpsiq.org). This article reports the main results of the first wave of COVIDPsiq study, which happened one month after the implementation of contagion measures by the Brazilian government. The aim of this study was to investigate and compare the independent effects of demographics, health-related anxiety, social distancing, exposure to the news of coronavirus pandemic, substance use, and traumatic situations on stress, anxiety, depression, and PTSD symptoms.

## 2. Methods

### 2.1 Study design

This is a cross-sectional study nested to a prospective cohort developed to enroll participants within nine months of follow-up during the COVID-19 outbreak, using a non-probabilistic, convenience sample. A complete description of the methodology of the longitudinal study is available elsewhere (Crestani Calegaro et al., 2020).

### 2.2 Participants and context

The inclusion criteria were to live in Brazil with 18 years or over. Participants with inconsistent answers and/or incomplete demographic questionnaire (minimal information) were excluded.

The COVIDPsiq study has begun collecting data from April 22^nd^ to May 8^th^, two months after the confirmation of the first case in São Paulo (February 26^th^, 2020), and 41 days after the Brazilian Health Ministry had declared community transmission (March 11^th^, 2020). At the end of this phase, Brazil had summed 145,328 cases and 9,897 deaths (Ministério da Saúde, 2020). Within the south region, where most of participants was located, the spread of contagion was still centered in metropolitan areas. Since the main portion of the sample was from the state of Rio Grande do Sul, we describe some measures adopted prior to the survey. In march 16^th^, public calamity was declared, and the first restrictions were implemented, for example: prohibition of interstate collective transportation, activities in shopping centers, beaches, and big events (Rio Grande Do Sul, 2020a). In April 1^st^, more restrict measures were adopted, prohibiting the operation of schools, universities and most commercial establishments, except essential services as drug stores, grocery stores, gas stations, and other (Rio Grande Do Sul, 2020b).

### 2.3 Measures

The study protocol included demographics, occupational status (including health professional and contact with public attendance), income and unemployment, infection by SARS-COV-2, COVID-19 in relatives and close people, information about emotional and sexual abuse, physical violence, psychiatric history, frequency, and intensity of exposure to news related to the pandemic, social distancing and isolation, and substance use. The Alcohol Use Disorders Identification Test - Consumption (AUDIT-C), was applied to measure the level of alcohol abuse. It is a three-item test derived from the complete assessment (AUDIT), particularly useful to identify heavy drinkers (Taufick et al., 2014). A score of 6-7 indicate high risk of dependence, and 8 or more, severe.

Two self-report instruments were used as main outcomes. The Depression, Anxiety and Stress Scale (DASS-21) is a 21-item *Likert* four point scale (0, 1, 2 and 3) that measures symptom severity of these three domains (Vignola and Tucci, 2014). DASS-21 is derived from the concept that stress is implicated in depression and anxiety, being a common component in both. Depressive symptoms are assessed as mild when the score on the items corresponding to depression is 10-13, moderate when 14-20, severe when 21-27 and extremely severe if greater than 27. Anxious symptoms are assessed as mild when the score in items corresponding to anxiety are 8-9, moderate when 10-14, severe when 15-19 and extremely severe if greater than 20. Stress symptoms are assessed as mild when the score on the corresponding items is 15-18, moderate when 19-25, severe when 26-33 and extremely severe if greater than 34 (Vignola and Tucci, 2014). The PTSD Checklist for DSM-5 (PCL-5) is a 20-item questionnaire widely used for screening and monitoring the severity of symptoms over time (Lima et al., 2016). A score of 38, out of a maximum of 80, is associated with the probable PTSD (Blevins et al., 2015).

### 2.4 Procedures

Data was gathered through a web-based survey using the SurveyMonkey® virtual platform. The engagement of participants was spontaneous, accounting with a snow-ball technique. Research disclosure included social media networks (Facebook®, Instagram®, Twitter® and LinkedIn®), corporative mailing list (emails sent by higher education institutions, governmental departments, and professional councils), digital and press media, cast news on radio and television, and through the most popular message applications in Brazil (WhatsApp® and Messenger®). The advertisements stated that the survey was anonymous, and participants had to give consent to take part in the study. The choice of an electronic survey was based on the possibility to reach more participants while respecting social isolation restrictions in Brazil.

### 2.5 Data analysis

The data has been stored on the SurveyMonkey® server, with access through the principal investigator’s (PI) account (VC). The database was anonymized and deidentified and can be accessed by request to the PI (VC). Data treatment and preparation (such as verification of normality and analysis of missing values) were performed. Continuous variables (age, AUDIT-C, DASS-21, and PCL-5) were not normally distributed; then, Mann-Whitney and Kruskal-Wallis (followed by pairwise comparisons) were preferred for bivariate tests, as well as Spearman’s correlations.

Multivariate analyses were performed through hierarchical multiple linear regression models (HMLR) to estimate the contribution of each block of independent variables on the outcomes (DASS-21 stress, anxiety and depression, and PCL-5 scores). For each dependent variable, we built five models, from low to high complexity, adding blocks of related variables, and testing prediction improvement (Kline, 2016). All blocks included variables using forced entry, as follows: 1) controlling variables (age, sex and education); 2) socioeconomic data; 3) variables related with the pandemic (social distancing, traumatic situations, symptoms of COVID-19, clinical comorbidities, and distress related to the pandemic news); 4) psychiatric information, treatment, and sleep quality; and 5) use of substances (alcohol, tobacco, and use of anxiolytics, opioids, cannabis, and stimulant drugs at least once a month). No substantial multicollinearity was identified, and residuals were normally distributed. Outliers were excluded from the analysis. A significance level of 5% was considered for all statistical tests. Statistical analyses were performed using SPSS© v23. Missingness were treated by pairwise and listwise methods in bivariate and multivariate analyses, respectively.

### 2.6 Ethical considerations

The study was approved by the National Research Ethics Committee (CONEP; CAAE: 30420620.5.0000.5346). All participants were voluntary and agreed with the Informed Consent Term. Contact information for mental health care available at the Brazilian Unified Health System (SUS) was provided at the end of the study protocol. Participants were also encouraged to contact the researchers in case of emotional discomfort through the Support to the Participant Service (SPS), available in the research website (www.covidpsiq.org/SAP). The SPS provided medical and psychological teleattendance by email and video calls, using psychoeducation and guidance to help participants to cope with stress and referring those with clinically relevant symptoms to mental health services.

## 3. Results

During the first wave of this study (T0), 3,796 answers were achieved. Of them, 164 entries were excluded due to duplicity or missing demographics. Brazilians who were abroad during the study period were excluded from the analysis (38). Seven cases were also excluded due to inconsistent and contradictory answers. A total of 3,587 participants were included. The sample composition is described at Table 1. Figure 1 shows the prevalence of psychiatric diagnoses.

**Table 1.** Sample characteristics.

|  |  | N | % |
| --- | --- | --- | --- |
| Sex | Male | 849 | 23.7% |
|  | Female | 2738 | 76.3% |
| Age (*Median; IQI*) |  | 29.0 | 19.0 |
| Education | Primary or secondary | 339 | 9.5% |
|  | Undergraduate | 1301 | 36.3% |
|  | Graduated | 665 | 18.5% |
|  | Post-graduated | 1282 | 35.7% |
| Ethnicity | White | 3069 | 85.6% |
|  | Non-white | 518 | 14.4% |
| Marital status | Single, divorced or widowed | 2274 | 63.4% |
|  | Married or stable relationship | 1313 | 36.6% |
| Living arrangement | With family or partner | 2647 | 73.8% |
|  | With other people | 413 | 11.5% |
|  | Alone | 527 | 14.7% |
| Occupation | Health worker | 653 | 18.2% |
|  | Worker | 1280 | 35.7% |
|  | Student | 1317 | 36.7% |
|  | Unemployed | 194 | 5.4% |
|  | Retired | 143 | 4.0% |
| Family income (minimum salaries<sup>a</sup>) | 1 or less | 231 | 6.4% |
|  | 1 to 2 | 457 | 12.7% |
|  | 2 to 8 | 1667 | 46.5% |
|  | 8 to 11 | 470 | 13.1% |
|  | 11 or more | 762 | 21.2% |
| Social distancing | No | 463 | 14.1% |
|  | Yes, alone | 328 | 10.0% |
|  | Yes, accompanied | 2486 | 75.9% |
| Financial indebtedness<sup>b</sup> | Improbable | 1226 | 37.4% |
|  | Possible | 1366 | 41.7% |
|  | Highly probable | 434 | 13.2% |
|  | Already in debt | 252 | 7.7% |
| Clinical comorbidity of risk | Any | 989 | 29.5% |
|  | *Diabetes mellitus* | 70 | 2.1% |
|  | High blood pressure | 237 | 7.1% |
|  | Respiratory problems | 402 | 12.0% |
|  | HIV/AIDS | 18 | 0.5% |
|  | Cancer | 41 | 1.2% |
|  | Cardiovascular problems | 61 | 1.8% |
|  | Obesity | 341 | 10.2% |
|  | Use of immune suppressor | 76 | 2.3% |
| COVID-19 | Flu-like syndrome<sup>c</sup> | 56 | 1.7% |
|  | Suspected or confirmed<sup>d</sup> | 60 | 1.8% |
| COVID-related trauma | Any | 200 | 6.1% |
|  | Living with person that had COVID-19 | 66 | 2.0% |
|  | Had distancing of close person due to COVID-19 | 54 | 1.6% |
|  | Someone close was hospitalized due to COVID-19 | 67 | 2.0% |
|  | Loss of relative due to COVID-19 | 17 | 0.5% |
|  | Loss of close person due to COVID-19 | 26 | 0.8% |
| Abuse/violence after the onset of pandemic | Any | 395 | 12.8% |
|  | Emotional abuse | 388 | 12.6% |
|  | Sexual abuse | 5 | 0.2% |
|  | Physical violence | 28 | 0.9% |
| Current psychological or psychiatric treatment | No | 2258 | 67.3% |
|  | Treatment maintained during pandemic | 576 | 17.2% |
|  | Treatment interrupted during pandemic | 523 | 15.6% |
| Substance use | Tobacco | 279 | 8.3% |
|  | Benzodiazepines<sup>e</sup> | 263 | 7.3% |
|  | Opioids<sup>e</sup> | 131 | 3.7% |
|  | Cannabis<sup>f</sup> | 446 | 13.3% |
|  | Cocaine, ecstasy or LSD<sup>f</sup> | 172 | 5.1% |
| Risk of alcohol dependence (AUDIT-C) | No risk/low | 1859 | 55.4% |
|  | Moderate | 1045 | 31.1% |
|  | High | 285 | 8.5% |
|  | Severe | 168 | 5.0% |
| Stress (DASS-21) | Normal | 1302 | 41.6% |
|  | Mild | 406 | 13.0% |
|  | Moderate | 553 | 17.7% |
|  | Severe | 531 | 17.0% |
|  | Extremely severe | 340 | 10.9% |
| Anxiety (DASS-21) | Normal | 1495 | 47.7% |
|  | Mild | 237 | 7.6% |
|  | Moderate | 569 | 18.2% |
|  | Severe | 269 | 8.6% |
|  | Extremely severe | 562 | 17.9% |
| Depression (DASS-21) | Normal | 1215 | 38.8% |
|  | Mild | 396 | 12.6% |
|  | Moderate | 641 | 20.5% |
|  | Severe | 337 | 10.8% |
|  | Extremely severe | 543 | 17.3% |
| PTSD (PCL-5) | Normal | 2271 | 75.5% |
|  | Probable PTSD | 736 | 24.5% |

Notes: <sup>a</sup> Minimum salary was R\$ 1,045.00 (corresponding to US\$ 192.17 in 04/30/2020). <sup>b</sup> Subjective perception of the probability of financial indebtedness. <sup>c</sup> Flu-like syndrome was considered when participants presented fever plus at least one of: cough, dyspnea, sore throat, or coryza. <sup>d</sup> Suspected COVID-19 according to a medical evaluation. Confirmed COVID-19 by laboratory tests. <sup>e</sup> Use of benzodiazepines or opioids (morphine and derivatives) at least once a month, without follow-up by a doctor. <sup>f</sup> Use of cannabis, cocaine, ecstasy or LSD at least once a month.

**Figure 1.**
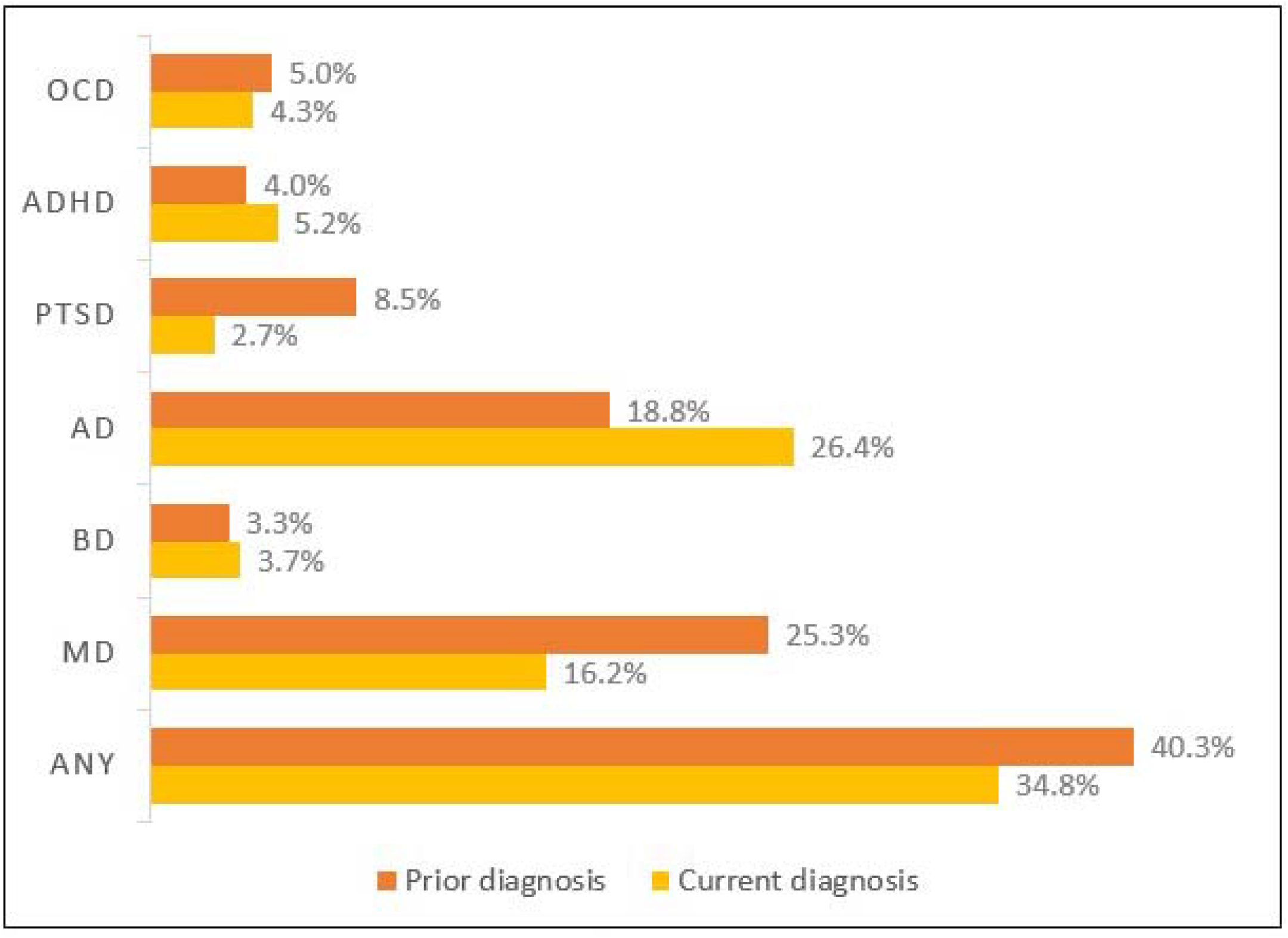
Prevalences of self-reported prior and current psychiatric diagnoses. Notes. OCD: obsessive-compulsive disoreder. ADHD: Attention deficit hyperactivity disorder. PTSD: post-traumatic stress disorder. AD: anxiety disorders (generalized anxiety disorder, social anxiety disorder, panic disorder). BD: bipolar disorder. MD: major depressive disorder.

Distribution across the country was heterogeneous, with 3163 (88.2%) respondents located in the southern region; 237 (6.6%), in the southeast; 187 (5,2%) in the other regions of Brazil. The city with more participants was Santa Maria (*n* = 1327; 37.0%), located at the center of the State of Rio Grande do Sul (*n* = 2698; 75.2%). Of the total (*n =* 3587), 3132 participants completed the DASS-21 scale, and 3007, the PCL-5 scale. Considering mild to extremely severe symptoms, the sample prevalence found was 58.4% for stress; 52.3% for anxiety; 61.2% for depression, and 24.5% for PTSD.

### 3.1 Bivariate analysis

Considering the inequity of the spread of coronavirus in Brazil, and the heterogeneity of the location of respondents, we begin by analyzing differences in the level of symptoms among regions, states, and cities. Despite of Kruskal-Wallis (K-W) test had shown significant differences in DASS-21 scales and PCL-5 among regions (*P*-values ranging from 0.017 to 0.048 from DASS-21 stress to PCL-5), these findings were not supported by pairwise comparisons. Moreover, K-W did not present significant differences in the main outcomes comparing the city where most of participants were in with other microregions, metropolitan areas, or regions, neither did comparing Rio Grande do Sul with other states.

Figure 2 shows that almost two thirds of the respondents considered their mental health had worsened after the social restriction measures initiated. The perception of change in mental health was correlated with DASS-21 stress (*rho* = −0.615), anxiety (*rho* = −0.531), depression (*rho* = −0.559), and PCL-5 (*rho* = −0.515). In other words, negative changes corresponded to high level of symptoms. All correlations were significant with *p* values *<* 0.001.

**Figure 2.**
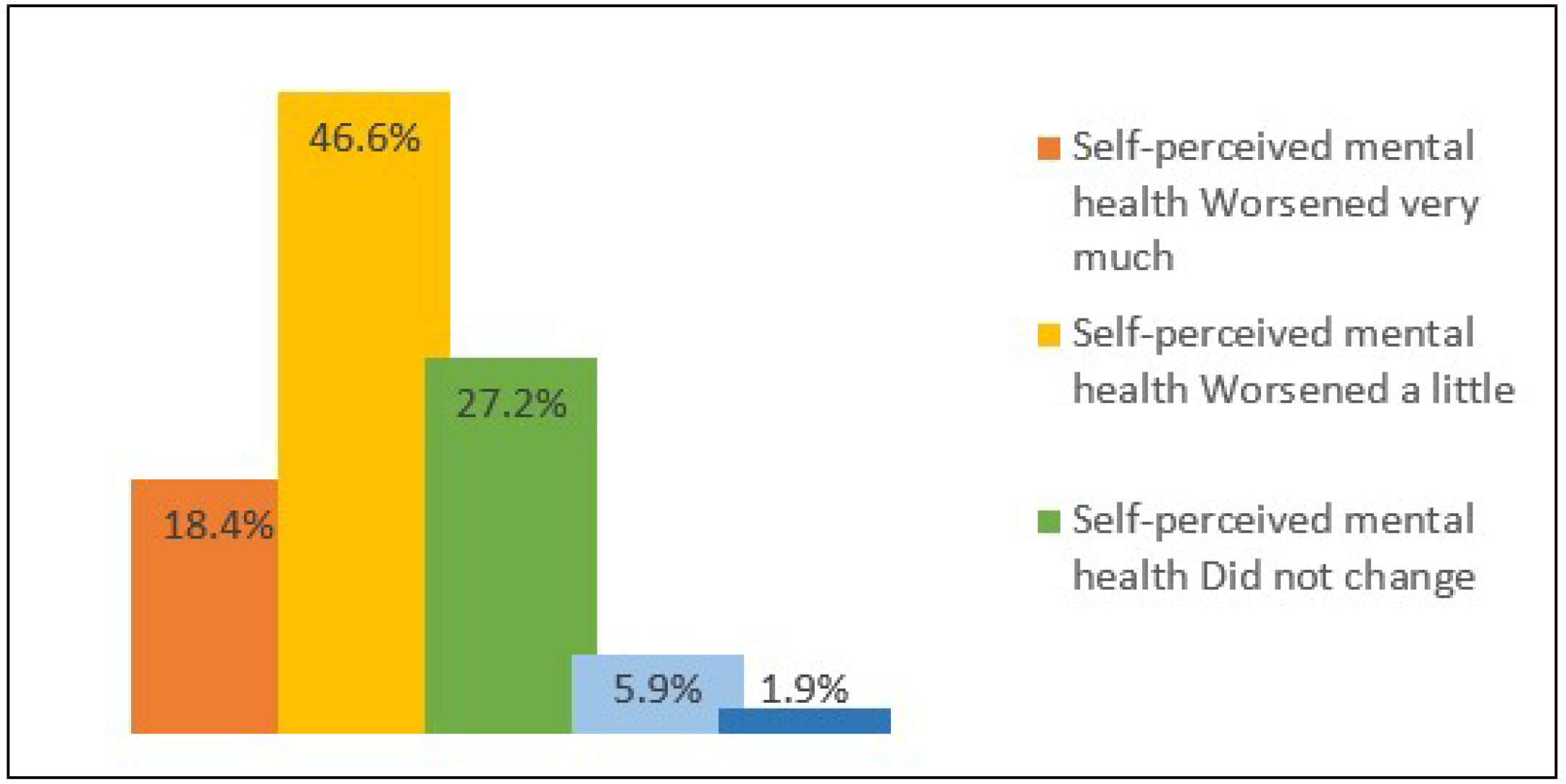
Self-perceived change in mental health after the onset of the pandemic.

Table 2 presents the results of M-W and K-W tests for bivariate analyses. Most variables were statistically associated with the outcomes, excepting diagnosis of COVID-19 suspected by a health professional or confirmed thru laboratory tests.

**Table 2.** Bivariate analysis of independent variables and symptoms of stress, anxiety, depression, and PTSD.

|  |  | DASS-21 Stress |  | DASS-21 Anxiety |  | DASS-21 Depression |  | PCL-5 |  |
| --- | --- | --- | --- | --- | --- | --- | --- | --- | --- |
|  |  | *Mdn*<br>[*IQI*] | *p* | *Mdn*<br>[*IQI*] | *p* | *Mdn*<br>[*IQI*] | *p* | *Mdn*<br>[*IQI*] | *p* |
| Sex | Male | 14 [16] | **< 0.001** | 4 [10] | **< 0.001** | 10 [14] | **< 0.001** | 15 [24] | **< 0.001** |
|  | Female | 20 [16] |  | 8 [12] |  | 14 [18] |  | 23 [27] |  |
| Age | 18 - 25 | 22 [16] | **< 0.001** | 10 [14] | **< 0.001** | 18 [20] | **< 0.001** | 28 [27] | **< 0.001** |
|  | 25 - 35 | 18 [16] |  | 8 [14] |  | 12 [16] |  | 21 [27] |  |
|  | 36 - 45 | 16 [12] |  | 6 [12] |  | 10 [14] |  | 18 [26] |  |
|  | 46 - 55 | 12 [12] |  | 4 [10] |  | 8 [12] |  | 13 [22] |  |
|  | 56 - 65 | 10 [10] |  | 2 [8] |  | 8 [12] |  | 10 [17] |  |
|  | 66 - 80 | 8 [10] |  | 4 [10] |  | 4 [10] |  | 9 [17] |  |
| Level of education | Primary or secondary | 20 [18] | **< 0.001** | 10 [16] | **< 0.001** | 16 [18] | **< 0.001** | 28 [27] | **< 0.001** |
|  | Undergraduate | 22 [18] |  | 10 [16] |  | 16 [20] |  | 27 [29] |  |
|  | Graduated | 16 [14] |  | 8 [12] |  | 12 [17] |  | 20 [26] |  |
|  | Post-graduated | 14 [14] |  | 6 [10] |  | 10 [12] |  | 16 [21] |  |
| Ethnicity | White | 18 [16] | **< 0.001** | 8 [14] | **< 0.001** | 12 [16] | **< 0.001** | 21 [27] | **< 0.001** |
|  | Non-white | 20 [18] |  | 10 [14] |  | 16 [18] |  | 27 [28] |  |
| Marital status | Single, divorced or widowed | 20 [16] | **< 0.001** | 8 [14] | **< 0.001** | 14 [16] | **< 0.001** | 25 [27] | **< 0.001** |
|  | Married or stable relationship | 14 [14] |  | 6 [10] |  | 8 [14] |  | 16 [25] |  |
| Occupation | Health worker | 14 [14] | **< 0.001** | 6 [12] | **< 0.001** | 8 [12] | **< 0.001** | 16 [23] | **< 0.001** |
|  | Worker | 16 [16] |  | 6 [12] |  | 10 [16] |  | 19 [25] |  |
|  | Student | 22 [16] |  | 10 [14] |  | 16 [20] |  | 27 [27] |  |
|  | Unemployed | 20 [16] |  | 10 [14] |  | 16 [20] |  | 29 [31] |  |
|  | Retired | 10 [14] |  | 4 [10] |  | 8 [14] |  | 10 [27] |  |
| Family income (minimum salaries) | 1 or less | 24 [16] | **< 0.001** | 14 [18] | **< 0.001** | 20 [20] | **< 0.001** | 33 [27] | **< 0.001** |
|  | 1 to 2 | 22 [16] |  | 12 [16] |  | 18 [20] |  | 31 [29] |  |
|  | 2 to 8 | 18 [16] |  | 8 [14] |  | 14 [16] |  | 23 [27] |  |
|  | 8 to 11 | 16 [16] |  | 6 [12] |  | 10 [14] |  | 19 [24] |  |
|  | 11 or more | 14 [16] |  | 4 [10] |  | 8 [13] |  | 14 [20] |  |
| Financial indebtedness | Improbable | 14 [16] | **< 0.001** | 6 [10] | **< 0.001** | 10 [16] | **< 0.001** | 17 [23] | **< 0.001** |
|  | Possible | 18 [16] |  | 8 [14] |  | 12 [16] |  | 22 [26] |  |
|  | Highly probable | 20 [16] |  | 10 [14] |  | 16 [18] |  | 28 [31] |  |
|  | Already in debt | 22 [18] |  | 12 [16] |  | 20 [20] |  | 32 [33] |  |
| Social distancing | No | 14 [14] | **< 0.001** | 6 [10] | **< 0.001** | 8 [14] | **< 0.001** | 17 [25] | **< 0.001** |
|  | Yes, alone | 16 [14] |  | 6 [14] |  | 14 [17] |  | 21 [29] |  |
|  | Yes, accompanied | 18 [16] |  | 8 [14] |  | 14 [18] |  | 22 [27] |  |
| Clinical diagnosis of risk | No | 18 [16] | **0.004** | 8 [12] | **< 0.001** | 12 [16] | **0.009** | 21 [26] | **0.001** |
|  | Yes | 18 [18] |  | 10 [16] |  | 14 [18] |  | 23 [30] |  |
| Flu-like syndrome | No | 18 [16] | 0.0814 | 8 [14] | **0.005** | 12 [16] | **0.008** | 21 [27] | **0.087** |
|  | Yes | 20 [18] |  | 14 [18] |  | 18 [20] |  | 26 [35] |  |
| Suspected or confirmed COVID-19 | No | 18 [16] | 0.956 | 8 [14] | 0.779 | 12 [16] | 0.518 | 21 [27] | 0.927 |
|  | Yes | 17 [13] |  | 10 [14] |  | 12 [16] |  | 21 [22] |  |
| COVID-related trauma | No | 18 [16] | **0.010** | 8 [14] | **0.001** | 12 [16] | **0.047** | 21 [27] | **0.005** |
|  | Yes | 20 [16] |  | 10 [16] |  | 14 [14] |  | 26 [27] |  |
| Intensity of media exposure | Mild | 16 [14] | **< 0.001** | 6 [10] | **< 0.001** | 10 [18] | **< 0.001** | 15 [24] | **< 0.001** |
|  | Moderate | 16 [14] |  | 6 [12] |  | 10 [16] |  | 20 [25] |  |
|  | Constant | 18 [16] |  | 8 [14] |  | 12 [16] |  | 22 [26] |  |
|  | Extreme | 20 [18] |  | 10 [14] |  | 16 [20] |  | 26 [31] |  |
| Emotional abuse/violence | No | 16 [14] | **< 0.001** | 6 [12] | **< 0.001** | 10 [16] | **< 0.001** | 19 [25] | **< 0.001** |
|  | Yes | 26 [14] |  | 14 [16] |  | 22 [20] |  | 36 [27] |  |
| Previous psychiatric diagnosis | No | 16 [18] | **< 0.001** | 6 [12] | **< 0.001** | 10 [18] | **< 0.001** | 19 [27] | **< 0.001** |
|  | Yes | 20 [14] |  | 10 [14] |  | 14 [18] |  | 25 [26] |  |
| Current psychiatric diagnosis | No | 14 [14] | **< 0.001** | 5 [10] | **< 0.001** | 10 [12] | **< 0.001** | 16 [22] | **< 0.001** |
|  | Yes | 24 [16] |  | 14 [14] |  | 20 [18] |  | 34 [27] |  |
| Mental health treatment | No | 16 [16] | **< 0.001** | 6 [12] | **< 0.001** | 10 [16] | **< 0.001** | 19 [25] | **< 0.001** |
|  | Maintained during pandemic | 20 [14] |  | 10 [14] |  | 14 [18] |  | 26 [26] |  |
|  | Interrupted during pandemic | 22 [16] |  | 12 [16] |  | 18 [18] |  | 29 [30] |  |
| Sleep duration | > 8 hours | 20 [16] | **< 0.001** | 8 [16] | **< 0.001** | 16 [20] | **< 0.001** | 25 [28] | **< 0.001** |
|  | 6 to 8 hours | 16 [16] |  | 6 [12] |  | 10 [14] |  | 18 [25] |  |
|  | < 6 hours | 24 [14] |  | 14 [16] |  | 16 [20] |  | 32 [30] |  |
| Sleep latency | < 20 minutes | 12 [14] | **< 0.001** | 4 [10] | **< 0.001** | 8 [14] | **< 0.001** | 13 [20] | **< 0.001** |
|  | 20 to 30 minutes | 16 [12] |  | 6 [10] |  | 10 [14] |  | 19 [23] |  |
|  | 30 to 60 minutes | 20 [16] |  | 10 [14] |  | 14 [16] |  | 26 [25] |  |
|  | > 60 minutes | 26 [14] |  | 14 [16] |  | 20 [20] |  | 37 [29] |  |
| Use of tobacco | No | 18 [16] | 0.700 | 8 [14] | 0.344 | 12 [16] | **0.011** | 21 [27] | **0.020** |
|  | Yes | 18 [20] |  | 8 [16] |  | 16 [20] |  | 25 [32] |  |
| Use of benzodiazepines | No | 16 [16] | **< 0.001** | 8 [12] | **< 0.001** | 12 [18] | **< 0.001** | 21 [27] | **< 0.001** |
|  | Yes | 24 [14] |  | 12 [14] |  | 18 [20] |  | 31 [28] |  |
| Use of opioids | No | 18 [16] | **0.003** | 8 [14] | **< 0.001** | 12 [16] | **0.003** | 21 [27] | **< 0.001** |
|  | Yes | 20 [16] |  | 12 [14] |  | 16 [20] |  | 28 [29] |  |
| Use of cannabis | No | 16 [16] | **< 0.001** | 6 [12] | **< 0.001** | 12 [18] | **< 0.001** | 20 [27] | **< 0.001** |
|  | Yes | 22 [16] |  | 12 [14] |  | 18 [18] |  | 28 [27] |  |
| Use of cocaine, ecstasy, or LSD | No | 18 [16] | **0.001** | 8 [14] | **0.006** | 12 [16] | **< 0.001** | 21 [27] | **< 0.001** |
|  | Yes | 22 [16] |  | 10 [16] |  | 16 [20] |  | 30 [31] |  |

Notes: *Mdn*: median. *IQI*: interquartile interval. DASS-21: Depression, Anxiety, and Stress Scale. PCL-5: Posttraumatic Checklist for DSM-5. LSD: lysergic acid. Significant values are highlighted in bold. Mann-Whitney and Kruskal-Wallis tests were used.

Continuous variables were analyzed through Spearman’s correlations. Age was negatively correlated with DASS-21 Stress (*rho* = −0.333; *p <* 0.001), Anxiety (*rho* = −0.273; *p <* 0.001), Depression (*rho* = −0.330; *p <* 0.001); and PCL-5 (*rho =* −0.276; *p <* 0.001). Intensity of distress related to pandemic news was correlated with DASS-21 Stress (*rho =* 0.531; *p* < 0.001), Anxiety (*rho =* 0.505; *p* < 0.001), Depression (*rho =* 0.446; *p <* 0.001); and PCL-5 (*rho =* 0.485; *p* < 0.001). AUDIT-C was weakly correlated with DASS-21 Stress (*rho* = 0.080; *p* < 0.001), Anxiety (*rho* = 0.044; *p* = 0.013), Depression (*rho* = 0.089; *p* < 0.001); and PCL (*rho* = 0.060; *p* = 0.001).

### 3.2 Multivariate analysis

Table 3 highlights the contribution of each block to explain the variance of the outcomes. In decrescent order, the block 3, which included variables related with the pandemic, was the most important. Then, block 1, composed by age, sex, and level of education, and block 4, psychiatric diagnoses, treatment, and sleep disturbances. The block 5 accounted for less than 1% of the total variance. All models were statistically significant.

**Table 3.** Hierarchical multiple linear regression model statistics.

| Outcome | Model | Block | Adjusted $R^2$ | $\Delta R^2$ | $F$ change | $fd1$ | $fd2$ | $p$ |
| --- | --- | --- | --- | --- | --- | --- | --- | --- |
| DASS-21 Stress | 1 | Age, sex, and education | 0.159 | 0.160 | 187.908 | 3 | 2966 | < **0.001** |
|  | 2 | Socioeconomic factors | 0.192 | 0.035 | 16.088 | 8 | 2958 | < **0.001** |
|  | 3 | COVID-related variables | 0.422 | 0.231 | 148.495 | 8 | 2950 | < **0.001** |
|  | 4 | Psychiatric characteristics | 0.499 | 0.078 | 65.851 | 7 | 2943 | < **0.001** |
|  | 5 | Substance use | 0.503 | 0.005 | 4.667 | 6 | 2937 | < **0.001** |
| DASS-21 Anxiety | 1 | Age, sex, and education | 0.122 | 0.123 | 138.175 | 3 | 2963 | < **0.001** |
|  | 2 | Socioeconomic factors | 0.165 | 0.046 | 20.265 | 8 | 2955 | < **0.001** |
|  | 3 | COVID-related variables | 0.356 | 0.191 | 110.169 | 8 | 2947 | < **0.001** |
|  | 4 | Psychiatric characteristics | 0.452 | 0.097 | 75.240 | 7 | 2940 | < **0.001** |
|  | 5 | Substance use | 0.454 | 0.003 | 2.552 | 6 | 2934 | **0.018** |
| DASS-21 Depression | 1 | Age, sex, and education | 0.120 | 0.121 | 136.115 | 3 | 2967 | < **0.001** |
|  | 2 | Socioeconomic factors | 0.176 | 0.058 | 25.959 | 8 | 2959 | < **0.001** |
|  | 3 | COVID-related variables | 0.347 | 0.173 | 98.227 | 8 | 2951 | < **0.001** |
|  | 4 | Psychiatric characteristics | 0.428 | 0.082 | 60.785 | 7 | 2944 | < **0.001** |
|  | 5 | Substance use | 0.435 | 0.007 | 6.464 | 6 | 2938 | < **0.001** |
| PCL-5 | 1 | Age, sex, and education | 0.111 | 0.112 | 120.044 | 3 | 2851 | < **0.001** |
|  | 2 | Socioeconomic factors | 0.177 | 0.068 | 29.555 | 8 | 2843 | < **0.001** |
|  | 3 | COVID-related variables | 0.372 | 0.196 | 111.603 | 8 | 2835 | < **0.001** |
|  | 4 | Psychiatric characteristics | 0.473 | 0.101 | 78.355 | 7 | 2828 | < **0.001** |
|  | 5 | Substance use | 0.477 | 0.005 | 4.284 | 6 | 2822 | < **0.001** |

Notes. DASS-21: Depression, Anxiety and Stress Scale. PCL-5: Posttraumatic Checklist for DSM-5. Statistically significant $p$ -values are highlighted in bold. $R^2$ represents the percentage of the outcome variable explained by the model. $\Delta R^2$ means the amount of improve in the prediction of the outcome when a new block of variables is added to the model. $F$ statistic indicates the ANOVA statistic for the model. $fd$ :: freedom degrees.

Table 4 presents the resulting coefficients of hierarchical multiple linear regression. In block 1, age was negatively associated with the levels of stress, anxiety, depression, and PTSD. Female sex predicted all outcomes except depression, and level of education only predicted anxiety. Note that these variables had higher size effects for stress and anxiety than depression and PTSD.

**Table 4.** Hierarchical multiple linear regression coefficients for DASS-21 Stress, Anxiety, and Depression, and PCL-5 scores.

|  | DASS-21 Stress |  | DASS-21 Anxiety |  | DASS-21 Depression |  | PCL-5 |  |
| --- | --- | --- | --- | --- | --- | --- | --- | --- |
| | $\beta$ | *p* | $\beta$ | *p* | $\beta$ | *p* | $\beta$ | *p* |
| Age | -0.245 | < **0.001** | -0.184 | < **0.001** | -0.181 | < **0.001** | -0.163 | < **0.001** |
| Female (vs. male) | 0.094 | < **0.001** | 0.075 | < **0.001** | 0.022 | 0.140 | 0.047 | **0.001** |
| Level of education | -0.015 | 0.357 | -0.072 | < **0.001** | -0.014 | 0.422 | -0.028 | 0.118 |
| Single, divorced, or widowed (vs. married) | 0.016 | 0.334 | 0.007 | 0.682 | -0.015 | 0.379 | -0.004 | 0.833 |
| Non-white ethnicity (vs. white) | 0.011 | 0.429 | 0.014 | 0.322 | 0.011 | 0.449 | 0.017 | 0.224 |
| Occupation - health worker<sup>a</sup> | 0.001 | 0.921 | 0.007 | 0.657 | -0.024 | 0.129 | -0.024 | 0.126 |
| Occupation – student<sup>a</sup> | 0.041 | **0.033** | -0.026 | 0.197 | 0.050 | **0.014** | 0.031 | 0.116 |
| Occupation – unemployed<sup>a</sup> | -0.008 | 0.592 | -0.008 | 0.570 | 0.032 | **0.031** | 0.020 | 0.170 |
| Occupation – retiree<sup>a</sup> | 0.018 | 0.236 | 0.037 | **0.021** | 0.034 | **0.035** | 0.032 | **0.046** |
| Family income | -0.019 | 0.211 | -0.076 | < **0.001** | -0.051 | **0.002** | -0.063 | < **0.001** |
| Financial indebtedness | 0.055 | < **0.001** | 0.037 | **0.011** | 0.074 | < **0.001** | 0.093 | < **0.001** |
| Social distancing - alone | 0.037 | **0.030** | 0.030 | 0.097 | 0.092 | < **0.001** | 0.057 | **0.002** |
| Social distancing - accompanied | 0.047 | **0.006** | 0.041 | **0.025** | 0.059 | **0.002** | 0.043 | **0.019** |
| Frequency of exposure to news of pandemic | 0.036 | **0.007** | 0.028 | **0.045** | 0.047 | **0.001** | 0.057 | < **0.001** |
| Intensity of distress related with news of pandemic | 0.352 | < **0.001** | 0.301 | < **0.001** | 0.259 | < **0.001** | 0.272 | < **0.001** |
| Clinical comorbidity of risk for COVID-19 | 0.036 | **0.007** | 0.052 | < **0.001** | 0.019 | 0.189 | 0.028 | **0.045** |
| Flu-like syndrome | 0.001 | 0.961 | 0.027 | 0.050 | 0.025 | 0.075 | 0.008 | 0.542 |
| COVID-related trauma | 0.005 | 0.690 | 0.028 | **0.047** | -0.004 | 0.765 | 0.007 | 0.630 |
| Emotional abuse/violence | 0.115 | < **0.001** | 0.081 | < **0.001** | 0.132 | < **0.001** | 0.148 | < **0.001** |
| Previous psychiatric diagnosis | 0.029 | **0.032** | 0.046 | **0.001** | 0.024 | 0.091 | 0.040 | **0.005** |
| Current psychiatric diagnosis | 0.160 | < **0.001** | 0.216 | < **0.001** | 0.174 | < **0.001** | 0.179 | < **0.001** |
| Maintained psychiatric treatment | 0.033 | **0.021** | -0.008 | 0.590 | 0.009 | 0.550 | 0.031 | **0.042** |
| Interrupted psychiatric treatment | 0.018 | 0.204 | 0.003 | 0.822 | 0.034 | **0.027** | 0.030 | **0.048** |
| Long sleep duration<sup>b</sup> | -0.019 | 0.170 | -0.015 | 0.308 | 0.081 | < **0.001** | -0.006 | 0.661 |
| Short sleep duration<sup>b</sup> | 0.095 | < **0.001** | 0.103 | < **0.001** | 0.052 | **0.001** | 0.081 | < **0.001** |
| Increased sleep latency | 0.156 | < **0.001** | 0.149 | < **0.001** | 0.158 | < **0.001** | 0.194 | < **0.001** |
| Use of tobacco | -0.009 | 0.511 | 0.001 | 0.958 | 0.037 | **0.012** | 0.030 | **0.037** |
| Alcohol consumption (AUDIT-C) | 0.050 | **0.001** | 0.016 | 0.302 | 0.038 | **0.016** | 0.032 | **0.041** |
| Use of benzodiazepines | 0.030 | **0.029** | 0.040 | **0.006** | 0.043 | **0.003** | 0.040 | **0.005** |
| Use of opioids | -0.021 | 0.110 | 0.008 | 0.567 | -0.013 | 0.350 | 0.008 | 0.589 |
| Use of cannabis | 0.026 | 0.090 | 0.021 | 0.192 | 0.036 | **0.031** | -0.016 | 0.336 |
| Use of cocaine, ecstasy, or LSD | -0.025 | 0.086 | -0.021 | 0.181 | -0.023 | 0.145 | 0.016 | 0.296 |

<sup>a</sup> Compared with workers. <sup>b</sup> Compared with sleep duration between 6 and 8 hours.
Notes: The different background colors represent each block in the hierarchical regression. Intercepts were omitted from the table. Coefficients are standardized $\beta$ s, which mean the amount of change in the outcome variable (in standard deviations) when the predictor increases one unit (or, in case of dichotomous variables, change from no to yes, for example). Standardized $\beta$ s are used to easily compare the effect size of each predictor. Statistically significant *p*-values are highlighted in bold.

On block 2, occupation was related with depressive symptoms, specifically being a student, unemployed or retiree. The last was also associated with anxiety and PTSD. Additionally, the greater the likelihood of indebtedness, greater psychological distress and the lower the household income, higher levels of anxiety, depression, and PTSD. Being a health worker did not increase the severity of symptoms at the time of this study. The same was true for ethnicity and marital status.

All the variables related more directly with the pandemic, included in block 3, were associated with the outcomes. The intensity of distress related with the pandemic news presented effect sizes 5 to 10 times greater than the frequency of exposition to information. Having a diagnosed clinical comorbidity of risk for COVID-19 increased anxiety, stress, and PTSD symptoms, whereas flu-like syndrome and COVID-related trauma were related only with increased anxiety. Emotional abuse or violence increased the severity of all symptoms, mostly depressive and post-traumatic stress. The social distancing was also associated with most outcomes, except the social distancing alone, which was not a predictor of anxious symptomatology. The symptoms of depression and PTSD were more influenced by being alone than being accompanied. The opposite has been found for stress and anxiety.

In block 4, psychiatric disorders and sleep disturbance were very important predictors of all dependent variables. Note that the current psychiatric diagnosis presented an effect size approximately 4 times greater than the previous psychiatric diagnosis. Also, ongoing or interrupted mental health treatment was associated with symptoms of post-traumatic stress. In addition, maintenance of treatment was associated with stress, while interruption was associated with depression. Short sleep duration and increased sleep latency were highly associated with the outcomes. The long sleep duration was more important than the short for predicting the symptoms of depression.

Finally, in block 5, the use of benzodiazepines without medical follow-up predicted all symptoms. The same was found for the level of alcohol consumption (AUDIT-C), except for anxiety. Moreover, tobacco use predicted symptoms of depression and PTSD, and cannabis, symptoms of depression.

## 4. Discussion

The current study presents the results of the baseline assessment of the COVIDPsiq study, through an internet-based survey applied approximately one month after the declaration of community transmission of corona virus and the implementation of measures to contain its spread. It must be interpreted considering a context in which participants were facing primarily the fear of the pandemic and the diverse consequences of the social distancing than infection, hospitalization, or loss of a close person due to COVID-19. Even though, two thirds of the participants declared worsening in their mental health condition. Despite the lack of an objective measure before the pandemic to document this change, the levels of stress, anxiety, depression, and PTSD were highly correlated with this self-perception. The main predictors of these symptoms were distress related with pandemic news, psychiatric diagnosis, lower age, sleeping problems, emotional abuse or violence, and economic problems, findings in accordance with previous reviews (Luo et al., 2020; Salari et al., 2020; Xiong et al., 2020).

The leading predictor of all outcomes was the level of distress related with the news of COVID-19. Findings indicate that subjective feelings raised by the information were more relevant than the frequency of exposure. Information and ways of communicating through technology may be pursued in order to cope with stress, as they enable maintenance of communication and social interaction, provide distraction and content, and can be a tool for education, work and information dissemination (Fineberg et al., 2018). However, excessive engagement in media and obsessive online activities may lead to severe problems, with significant risk of disordered and addictive use (Vismara et al., 2020). Excessive media exposure is an impairing repetitive behavior highly associated with distress intensity and negative outcomes. Gao et al (2020) showed an association of high media exposure with increased risk of anxiety and comorbid major depressive disorder compared with low exposure during COVID-19 outbreak in Wuhan, China (Gao et al., 2020). As referred by the World Health Organization (WHO), the infodemic phenomenon can difficult searching for trustworthy sources and leads to misinformation and information overload, which contributes to distress (World Health Organization, 2020).

We also suggest that the influence of the news on symptoms is linked to the way disasters are reported by the media. Although the coverage of this type of event is “emotional by nature, whether it focuses on the emotions of individuals directly affected by the tragic events or the collective emotions of the larger community reacting to the misfortunes of others like them” (Pantti and Wahl-Jorgensen, 2007), this aspect can be reinforced according to the used narrative strategies. The transmission of “faces, gestures, proper names, expressions of pain, images of victims (…) give a human and personal character to all catastrophes” (Amaral and Ascencio, 2015). At the same time, they cooperate to establish a public identification process with the suffering portrayed by the media. Thus, even the ones who were not directly affected by the disease imagine that they could have been victims. This is one of the pillars of the “virtual victim policy”, a narrative strategy based on the combination of suffering, fear and risk, in which the audience is encouraged to think that “the event might happen to any individual, it may happen again and it could have been avoided (Vaz et al., 2013).

Another concern pointed in this study was the vulnerability of people with psychiatric diagnosis to aggravate their symptoms. At the period that data was gathered, many public mental health outpatient clinics were operating with closed doors, offering telephone guidance for those seeking care. The interruption of treatment, as shown in the results, predicted levels of symptoms of depression and PTSD. In the absence of a doctor, people may use benzodiazepines, alcohol, and other drugs to cope with stress, anxiety, and insomnia, leading to more depression, chronic PTSD, substance dependence, and domestic violence (Telles et al., 2020).

In this line, sleep disruption was clearly associated with the symptoms of stress, anxiety, depression, and PTSD. It may raise due to the confinement of unknown duration, and/or fear of getting infected, as well when facing traumatic situations. Beck et al. (2020) conducted a cross-sectional study with a representative sample of the general population in France, and showed an increase of 25% of trouble sleeping comparing with a prior general population survey (performed in 2017) (Beck et al., 2020). In a retrospective survey conducted in China, Li et al. (2020) found that the prevalence of insomnia significantly increased after the COVID-19 outbreak, with 13.6% of participants developing new-onset, and 12.5%, worsening previous sleeping problems (Li et al., 2020). Furthermore, they also observed a significant increase of length of time in bed and total sleep time, and a decrease in sleep efficiency. Delayed bedtime and wakeup time were also noted. Studies have reported sleeping problems during pandemic more frequently among women, young adults, unemployed, people with financial problems, mental illness, media overexposure, COVID-19 related stress, increased severity of anxiety and depressive symptoms, and prolonged time in bed (Beck et al., 2020; Léger et al., 2020; Li et al., 2020). Daytime impairment and use of sleeping pills are usual consequences of trouble sleeping, increasing the potential for drug abuse and dependence (for example, alcohol or benzodiazepines). Such results are strongly consistent with those in the present study, which found that short sleep duration and increased sleep latency were among the most important predictors of stress, anxiety, depression, and PTSD, and long sleep duration predicting symptoms of depression.

Social distancing was independently associated with psychological distress, and we observed some differences among those alone comparing to those accompanied. Loneliness exerted more impact on depressive and post-traumatic stress symptoms, which can be explained by the deprivation of relationships, while to be accompanied was more relevant than to be alone regarding stress. Prati and Mancini (2021), meta-analyzed 25 recent studies on the impact of the COVID-19 pandemic on population mental health, and concluded that the psychological impact of social restrictions is small in magnitude and highly heterogeneous (Prati and Mancini, 2021). In our study, in fact social distancing played a secondary role to the explanation of the symptoms comparing the effect sizes with the main predictors, but the containment measures applied may had indirect effects, which were measured by other variables, i.e., economic problems and emotional abuse. For some people, staying more time with relatives or partner may flare interpersonal tension, disagreements, and, in some cases, family violence. The last refers to threats or other forms of violent behavior in families, which can be physical, sexual, psychological, or economic, in addition to child abuse and intimate partner violence. A raise in this behavior due to forced coexistence, economic stress, and fears about the corona virus has been observed in many countries (Bright et al., 2020; Campbell, 2020). In Brazil, between March 1^st^ to 25^th^, 2020, there was an 18% increase in the number of complaints of domestic violence produced by the “call 100” and “call 1808” services recorded by the National Ombudsman for Human Rights (ONDH) from the Ministry of Women, Family and Human Rights (MMFDH) (Vieira et al., 2020). According to Marques et al. (2020), during the pandemic, this type of violence is associated with an increase in the level of stress generated by the fear of falling ill, uncertainty about the future, increased consumption of alcohol and other psychoactive substances (Marques et al., 2020). Additionally, Vieira et al (2020) included economic stress, greater exposure to explorers, and reduced support options, such as contact with other relatives and friends, as risk factors (Vieira et al., 2020).

Socioeconomic factors were independently associated with many symptoms, pointing to potential sources of worries, frustration, and distress. Unemployment and financial indebtedness tend to worse during pandemics, which, in addition to low family income, can have lasting effects on people’s lives. According to the Well Being Trust, it is expected an increase on deaths of despair due to alcohol, drugs, and suicide, as consequence of unemployment in the years following the outbreak of the pandemic (Peterson et al., 2020). Furthermore, our findings showed that retirees presented more symptoms of anxiety, depression, and PTSD. Independently of age, retired either due to elderly or chronic disease may deal with reduced autonomy, financial problems, free time but deprivation of usual activities, thus becoming distressed and fearing the illness and the death. Of note, students presented higher levels of depression. They represented one-third of this sample and were mostly undergraduate or postgraduate studying remotely. Accordingly, a study carried out in Jordan during the Covid-19 pandemic by Naser et al. (2020) reported a higher prevalence of anxiety among university students (21.5%), comparing with health professionals (11.3%) and the general population (8.8%) (Naser et al., 2020). Before pandemic, Auerbach et al. (2016) had found an one-year prevalence of mental disorders in one-fifth of university students from 21 low to high-income countries (Auerbach et al., 2016). In this phase of life, young adults usually must cope with high academic demands, social pressure related to the future profession, and financial dependence. We hypothesize that concerns about possible impairments on practical learning, the lack of resources to access online classes, routine disruption, and the distancing from family, friends, and loved ones may raise feelings of sadness, inability, low self-esteem, abandonment, and hopelessness. In this sense, Wu et al. (2020) have shown that the effect of uncertainty stress on mental disorders were much superior than life and study stress, in a nationwide study with college students (Wu et al., 2020).

Having at least one comorbidity of risk for complications of COVID-19 predicted stress and PTSD, besides of anxiety symptoms. Additionally, flu-like syndrome and COVID-related trauma predicted symptoms of anxiety. Noteworthy, diagnosed, or suspected COVID-19 was not associated with the outcomes. Taken together, these issues suggest the fear of contagion, despite the disease itself. Note that at the time that data was gathered cases were emerging in Brazil, but most of the sample had not become infected or had lost a relative or friend. These findings can be explained by health anxiety, which may be associated with cyberchondria, and worsen with media overexposure (Jungmann and Witthöft, 2020). Health anxiety promotes preventive behaviors, such as washing hands and adhering to social distancing, but it also has a negative impact on work and family involvement (Ornell et al., 2020; Trougakos et al., 2020).

Demographics also played an important role in predicting the outcomes. The study found that the younger, the higher the levels of symptoms, with the effects of the age being more notable in stress. This group may suffer more with the distance of friends, family, loved ones, lack of social activities, job instability (Xiong et al., 2020). Although both sexes increased their psychological symptoms during the quarantine, women showed higher levels of stress, depression and anxiety (Jia et al., 2020). In agreement, other studies also found sex differences in the symptom distribution, with women reporting significantly higher levels of anxiety, depression (Solomou and Constantinidou, 2020) and PTSD (Liu et al., 2020) than men. Despite of the biological influence of sex, another hypothesis is the need to reconcile work with the care of children and domestic chores, which affects more the career progression and the salary of women than men. Finally, we found that low level of education was associated with symptoms of anxiety. In opposite way, the review of Salari et al. (2020), showed that higher levels of education were related to greater symptoms of stress, anxiety, and depression. The authors argued that the high level of education may lead to self-awareness of health, and therefore, to health anxiety. In Brazil, low education is linked to unemployment, poor working conditions, and low access to health services that may cause worry and anxiety.

This study must be understood considering its limitations. First, this is an internet-based survey in which convenience sample bias may limit its external validity. It does not represent northern regions of the country and, as several surveys of similar method, most participants are white wealthier females. Hence, extrapolations of the findings to other sociodemographic characteristics may not apply. Nonetheless, the sample is from an underrepresented area of the world, in a country with an important impact of the pandemic, which will be useful to compose the assessment of the global mental health outcomes of SARS-CoV-2. Second, all the pre-pandemic information was collected retrospectively. We attempt to minimize this by asking questions regarding current information, such as employment, living conditions and diagnosis, that are not likely be misled by recall bias. Third, it was not possible to evaluate if symptoms are clinically relevant to infer an increase incidence of mental illness. The symptomatic assessment utility lies on assessment of distress oscillation and comparability across the globe, using instruments validated in similar samples and widely used in other studies on the same topic.

Lastly, our current findings were able to put some light on relevant variables that, at the initial months of COVID-19 pandemic, appear to have a crucial role on symptoms of anxiety, depression, stress, and post-traumatic stress. It was shown that self-perception of suffering was highly correlated to the outcomes in study; self-referred level of distress related with the news of pandemic was the most important predictor of mental soreness of all, what may indicate that subjective issues are a large part of what compounds mental health symptoms during traumatic exposure. We identified other external and internal variables that were shown to have association with symptoms of anxiety, depression, stress, and PTSD, as age, psychiatric diagnosis, sleeping problems, emotional abuse and violence, female gender, being a student, retiree or unemployed, low income and probability of financial loss during quarantine, as well as substance use. These findings support the importance of some actions that may lead to a better event-coping strategy, as maintaining the operation of public healthcare institutions, so all layers of the population can have worthy access and attendance, mostly those who are already mentally ill. To avoid the risk of contagion and, at the same time, providing mental health support, telepsychiatry and telepsychotherapy are valuable solutions that may help patients with variable levels of severity (Kalin et al., 2020; Salum et al., 2020a, 2020b). The feasibility of online interventions, however, is dependent of technological resources, as broadband internet connections, availability of appropriate smartphones or computers, as well as ability to use them. This applies for both patients and care providers, requires investment and time to implement. Nevertheless, the economic and technologic inequities must be taken into account, as they limit the access of online interventions for elder, children, people of low-income, with disabilities and with cognitive impairment (Nadkarni et al., 2020). As a public health perspective, we strongly recommend that mental health services should receive investments to implement telehealth and to adapt the setting to meet biosafety regulations, to keep face-to-face attendance for those people without digital accessibility.

Considering that self-referred level of distress related with media exposure was the most significant predictor of mental outcome, it’s mandatory to considerer controlling strategies against compulsive use of technology as a matter of mental coping (Király et al., 2020). By the other hand, media can be a powerful ally to spread psychoeducational information, using many digital resources, such as videos, podcasts, and written material. They may be useful to clarify the risks of substance misuse, providing information on sleep hygiene, and encouraging people to seek appropriate treatment when necessary, indicating how to access and where to find mental health services of reference for example. Moreover, attention should be paid at specific stratum of our society, as students and those at risk of financial loss during the COVID19 pandemic; some actions as financial-security loans, flexibilization of study chronogram and active search of these individuals by mental health programs should lead to a better mental health scenario in the future.

## Data Availability

Data was gathered through a web-based survey using the SurveyMonkey® virtual platform. The results are available at the research website.

https://www.covidpsiq.org/

## Acknowledge

Dr. Hoffmann is supported by the research grant of the Brazilian Ministry of Health under the “Termo De Execução Descentralizada - TED 12/2019”.

